# Clinician contributions to disparities in severity of illness trajectories among mechanically ventilated patients

**DOI:** 10.64898/2026.06.23.26356358

**Authors:** Christopher F. Chesley, Olga Yakusheva, Yingying Lu, Rachel Kohn, Aerielle Belk, Stefania Scott, Scott D. Halpern, Meeta Prasad Kerlin

## Abstract

**Rationale:** Racial disparities in outcomes among patients with acute respiratory failure are well-described, but the contributions of clinicians to these disparities have not been evaluated.

**Objectives:** Among mechanically ventilated patients, we evaluated racial disparities in severity of illness trajectories and adapted value-added modeling to quantify nurse and physician relationships with these disparities.

**Methods:** In a retrospective cohort of mechanically ventilated patients across five hospitals between 2018 and 2022, we used generalized estimating equations to model the change in Laboratory-based Acute Physiology Score version 2 (LAPS) from the start to end of intensive care unit admission (Δ*LAPS*). Consistent with value-added modeling, we randomly allocated the cohort into development and testing partitions, and fit separate multiple linear regression models of Δ*LAPS* using concurrent nurse and physician assignments (determined at 4-hour intervals), patient race, and clinician-race interaction terms as fixed effects. Clinician-specific and clinician-race interaction coefficients were extracted to determine race-specific value-add for each clinician. We defined the race-contextual value-add difference (*RCVAD*) as a clinician-level measurement of the difference in that clinician’s value-add between Black and White patients in their care; a positive *RCVAD* indicates a more favorable severity of illness trajectory for Black relative to White patients and vice versa.

**Measurement and Main Results:** Among 6,555 distinct patients, 7,247 clinical encounters, 405 nurses, and 70 physicians, Black patients accounted for 2,926 (40%) encounters. Overall, Black patients had significantly less improvement in Δ*LAPS* than White patients (difference in LAPS decline = 2.26 [0.23, 4.29], p=0.029). In the development partition, median nurse *RCVAD* was −0.10 (interquartile range [IQR]: −1.17, 1.14) with 191 (47%) nurses having a positive *RCVAD*; median physician *RCVAD* was - 0.18 (IQR: −1.34, 0.56) with 29 (41%) having a positive *RCVAD*.

**Conclusions:** Black mechanically ventilated patients experience less improvement in severity of illness during intensive care unit admission than White patients. While the majority of physicians and nurses were associated with disparities-exacerbating illness trajectories, many other clinicians were associated with disparities-mitigating trajectories. Future work to understand practices associated with disparities-exacerbating and disparities-mitigating care profiles could inform interventions to reduce disparities overall.

## Introduction

Over 1.6 million Americans experience mechanical ventilation annually, with Black patients experiencing the highest risks of respiratory failure and mortality.^1–4^ Racial differences related to severity of illness at presentation may in part account for excessive mortality in Black compared to White mechanically ventilated patients.^5,6^ However, because racial disparities in clinical outcomes vary widely between centers,^3,5^ identifying clinical delivery factors associated with favorable severity of illness trajectories may be a strategy for developing interventions to mitigate disparities.

Value-added modeling is an econometric method that quantifies worker contributions to changes in a work-related quality measure. Applied to a clinical context, value-added modeling has quantified associations between the performance of individual nurses and improvements in severity of illness during hospital admissions.^7–9^ Recently, we applied value-added modeling in a two-clinician care delivery model consisting of nurses and physicians caring for mechanically ventilated patients, and demonstrated that individual nurse and physician care both independently contribute to severity of illness trajectory.^10^ In this study, we sought to adapt value-added modeling to evaluate race-specific differences in clinician performance. Our objectives were to 1) assess for independent relationships between patient race and severity of illness trajectories, and 2) adapt value-added modeling to quantify individual clinician-level differences in performance associated with patient race in a care delivery model consisting of nurses and physicians. We also sought to determine whether our approach satisfies identifying assumptions of conventional value-added modeling that mitigate potential biases associated with the method.^8,11^

## Methods

This retrospective cohort study was approved by the institutional review board at the University of Pennsylvania. We follow the Strengthening the Reporting of Observational Studies in Epidemiology (STROBE) reporting guidelines. Analyses were performed between winter 2025 and spring 2026.

### Study sites and participants

We included adult patients ≥18 years who received ≥12 hours of mechanical ventilation and who were admitted to one of six medical or mixed medical-surgical ICUs across five hospitals in the University of Pennsylvania Health System between April 1, 2018 and October 31, 2022. Only the first eligible episode was included for each patient during the same hospital encounter, but multiple hospital admissions for an individual patient were included if they contained a new eligible mechanical ventilation episode. Patients were excluded if they had orders for comfort care at the time of ICU admission or if admission age was recorded as >110 years, which likely reflected an indeterminable age for patients presenting without identifying information. Additionally, to allow for sample sizes that ensured stability of statistical models, only Black and White patients and only clinicians who provided care to both Black and White patients across ≥20 four-hour intervals were included.

### Data source and study variables

We extracted routinely collected clinical data maintained in the integrated electronic health record of the study health system. The outcome was the change in severity of illness from first day of mechanical ventilation in the ICU to the last day of ICU admission as determined by the Laboratory-based Acute Physiology Score, version 2 (Δ*LAPS*), an illness severity score that predicts inpatient mortality risk using physiologic and laboratory test variables.^12–14^ LAPS ranges from 0 to 256, with higher scores associated with higher mortality risk. Therefore, a positive Δ*LAPS* indicates a higher score at the end compared to the beginning of ICU admission and indicates a poor illness trajectory. Each daily Laboratory Acute Physiology Score (LAPS) was calculated using the most extreme value observed in the 24 hours preceding the day of record.^15^

Nurse and attending physician assignment were defined based on data in the electronic health record (EHR) for each four-hour interval from the start of mechanical ventilation to the end of the ICU admission. Patient race was defined as listed in the EHR.^10^ Potential confounders included patient age, sex, admission Elixhauser comorbidity index, admission source (emergency department, direct admission, or outside hospital transfer), medical vs surgical admission, hospital duration prior to mechanical ventilation initiation, interval day of the week, interval occurrence during the day (7am-6:59pm) or night (7pm-6:59am), and LAPS score on admission to the hospital.

### Statistical analysis

Statistical significance for all two-sided hypothesis tests was considered at p<0.05. Analyses were performed using Stata 18 (StataCorp, College Station, Texas, United States).

#### Model specification

We quantified relationships between patient race and Δ*LAPS* using a generalized estimating equation that regressed Δ*LAPS* on to patient race and the previously mentioned confounders with clustering based on study ICU. This model was performed at the encounter level and used a Gaussian distribution with identity link.

To estimate relationships between clinician assignment and race-specific severity of illness trajectory we adapted value-added modeling strategy. We developed a clinician value-add determination model at the level of the encounter-interval consisting of multivariable linear regression of Δ*LAPS* on to nurse and physician assignment (defined at four-hour intervals and included as separate fixed effects); patient race (a binary categorical variable, with White patients assigned the reference); patient race-clinician assignment interaction terms; and confounding variables as mentioned previously (including study ICU as a fixed adjustment variable).

#### Definition of race-contextual value-add difference

We extracted clinician 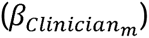 and race 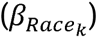 coefficients to define clinician value-add 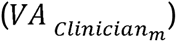 based on the following equation:

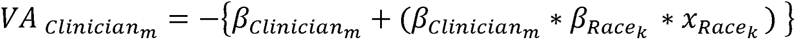

Where clinician *m*=1, 2, … refers to either nurse *i*=1, 2, … or physician *j*=1,2, …, and k=0 or 1 for White or Black patients, respectively. The expression is multiplied by negative one to preserve conventional interpretation of value-add estimates in which higher (more positive) *VA_Clinician_* is associated with more favorable severity of illness trajectory—a necessary step because higher LAPS is associated with greater mortality risk. Additionally, because White patients serve as the reference, 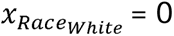 and 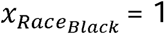.

*VA_Clinician_* was then standardized according to the following equation:

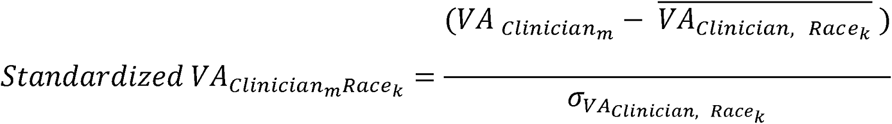

Where 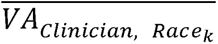 represents the race-specific mean value-add stratified by clinician group and 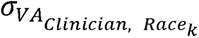 represents the race-specific standard deviation of the value-add estimate of the clinician group to which clinician *m* is a member. This standardization allows a relative comparison of value-add estimates between races in terms of their placement along a normal distribution stratified by race and specific to both clinician types. We then defined the race-contextualized value-add difference (*RCVAD*) for clinician *m* as:

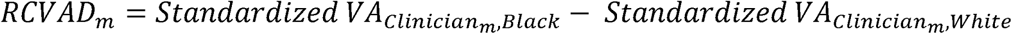

Thus, any clinician with a positive *RCVAD* is associated, on average, with a more favorable Δ*LAPS* for Black compared to White patients, and vice versa.

#### Evaluation of identifying assumptions

Conventional value-added modeling requires evaluating whether clinician-level value-add satisfies two identifying assumptions that reduce the potential for biased results:^8,10^ i) correlation of value-add estimates across random partitions of the study population and ii) random association of clinician value-add estimates with presenting patient severity of illness. To address the first assumption, we performed a 1:1 random split of all qualifying patient encounters into the development and testing partitions and determined the Pearson correlation (*r)* of clinician *RCVAD* estimates derived from both partitions. To test the second assumption, we first determined – for each clinician-encounter pair – a patient’s starting LAPS at the time of first assignment for each nurse-encounter or physician-encounter pair. Second, we determined the mean initial LAPS stratified by race for each clinician. Third, we calculated the difference between Black and White mean initial LAPS for each clinician. Finally, we assessed the correlation of the mean racial difference in initial LAPS and the *RCVAD* for each clinician between the two partitions.

## Results

The cohort included 6,555 patients with 7,247 distinct encounters, assigned to 679 nurses and 70 physicians. Black patients accounted for 2,569 (39%) patients and 2,926 (40%) encounters. This represented a total of 331,608 four-hour intervals, of which 283,470 (85%) were assigned a nurse, and 282,791 (85%) were assigned a physician after excluding clinicians based on exclusion criteria. Patient encounters were partitioned into the development (n=3,612) and testing (n=3,635) partitions. Within partitions, Black patients had higher admission LAPS, similar Δ*LAPS*, younger age, larger proportion of medical admissions, greater proportion of emergency department admissions, and longer duration of mechanical ventilation compared to White patients. Between partitions, between-race covariate balance was largely similar, including with respect to the median Δ*LAPS*, patient age, admission type, patient sex, admission source, admission LAPS, and number of clinician assignments. However, there were some differences between partitions, with Black patients having higher Elixhauser comorbidity indices and a similar hospitalization duration prior to mechanical ventilation relative to White patients in the development compared to the testing partition. Further baseline characteristics can be seen in **Table 1**.

**Table 1.** Encounter-level baseline study population characteristics by race and partition. Abbreviations: IQR, Interquartile range; ΔLAPS, change in Laboratory Acute Physiology Score (LAPS) from beginning to end of ICU discharge.

|  | Development partition |  | Testing partition |  |
| --- | --- | --- | --- | --- |
|  | White | Black | White | Black |
| Number of encounters, n (%) | 2145 (59%) | 1467 (41%) | 2176 (60%) | 1459 (40%) |
| ΔLAPS, median (IQR) | -30 (-57, -4) | -29 (-58, -3) | -31 (-59, -2) | -30 (-58, -2) |
| Patient age, median (IQR) | 65 (54, 74) | 62 (52, 71) | 65 (54, 74) | 63 (52, 72) |
| Admission type, n (%) |  |  |  |  |
| Medical admissions, n (%) | 1401 (65.3%) | 1110 (75.7%) | 1449 (66.6%) | 1104 (75.7%) |
| Surgical admissions, n (%) | 743 (34.6%) | 357 (24.3%) | 726 (33.4%) | 351 (24.1%) |
| Admission uncategorized, n (%) | 1 (<1%) | 0 (0.0%) | 1 (<1%) | 4 (0.3%) |
| Female sex, n (%) | 927 (43.2%) | 801 (54.6%) | 929 (42.7%) | 776 (53.2%) |
| Ethnicity, n (%) |  |  |  |  |
| Non-Hispanic | 2064 (96.2%) | 1429 (97.4%) | 2079 (95.5%) | 1431 (98.1%) |
| Hispanic | 48 (2.2%) | 25 (1.7%) | 62 (2.8%) | 16 (1.1%) |
| Unknown | 33 (1.5%) | 13 (0.9%) | 35 (1.6%) | 12 (0.8%) |
| Admission source, n (%) |  |  |  |  |
|  | White | Black | White | Black |
| Emergency department | 1170 (54.6%) | 1026 (70.0%) | 1179 (54.3%) | 1056 (72.5%) |
| Direct | 336 (15.7%) | 135 (9.2%) | 349 (16.1%) | 143 (9.8%) |
| Outside facility | 637 (29.7%) | 305 (20.8%) | 645 (29.7%) | 258 (17.7%) |
| Admission Elixhauser mortality index, median (IQR) | 21 (6, 36) | 23 (8, 38) | 22 (6, 36) | 22 (7, 38) |
| Admission LAPS, mean (SD) | 141 (49.3) | 148 (48.9) | 142(49.7) | 151 (49.7) |
| Hospitalization duration prior to mechanical ventilation, median hours (IQR) | 11.9 (3.4, 79) | 8.1 (4, 49.5) | 14.1 (3.8, 86.2) | 7.3 (3.7, 40.1) |
| Duration of mechanical ventilation, median hours (IQR) | 69.4 (31.3, 163.6) | 74.4 (35.0, 183.1) | 66.9 (30.3, 161.5) | 75.6 (35.2, 183.0) |
| Unassigned nurse assignments, n (%) | 11,456 (12%) | 12,332 (17%) | 11,819 (12%) | 12,531 (18%) |
| Unassigned physician assignments, n (%) | 17,422 (18%) | 8,051 (11%) | 14,938 (16%) | 8,406 (12%) |

### Associations between severity of illness trajectory and patient race

In unadjusted analysis, severity of illness trajectory measured by Δ*LAPS* was not significantly different between Black and White patients (mean difference 0.51[-1.62, 1.64], p=0.639). However, adjusted analyses demonstrated that Black patients experienced a more positive Δ*LAPS* relative to White patients (+2.26 [0.23, 4.29], p=0.029, **Table S1**). This suggests that Black patients experienced less improvement in severity of illness during ICU admission than did White patients.

### Associations between clinician value-add and racial differences in severity of illness trajectory

In the development partition, median nurse *RCVAD* was −0.10 (interquartile range: - 1.17, 1.14); 191 (47%) nurses had a positive *RCVAD*, indicating that just under half of nurses were associated with a relatively more favorable severity of illness change for Black relative to White patients on average. For physicians, median *RCVAD* was −0.18 (IQR: −0.95, 0.68); 29 (41%) physicians had a positive *RCVAD*. In the testing partition, median nurse *RCVAD* was 0.12 (IQR: −6.69, 4.36); 226 (56%) nurses had a positive *RCVAD*. Median physician *RCVAD* was −0.07 (IQR: −0.95, 0.68); 31 (44%) having a positive *RCVAD* (**Figure 1**). Though for each clinician type, median *RCVAD* was not identical across partitions, the minima, interquartile ranges, and maxima were comparable across partitions (**Table 2**).

**Figure 1A.**
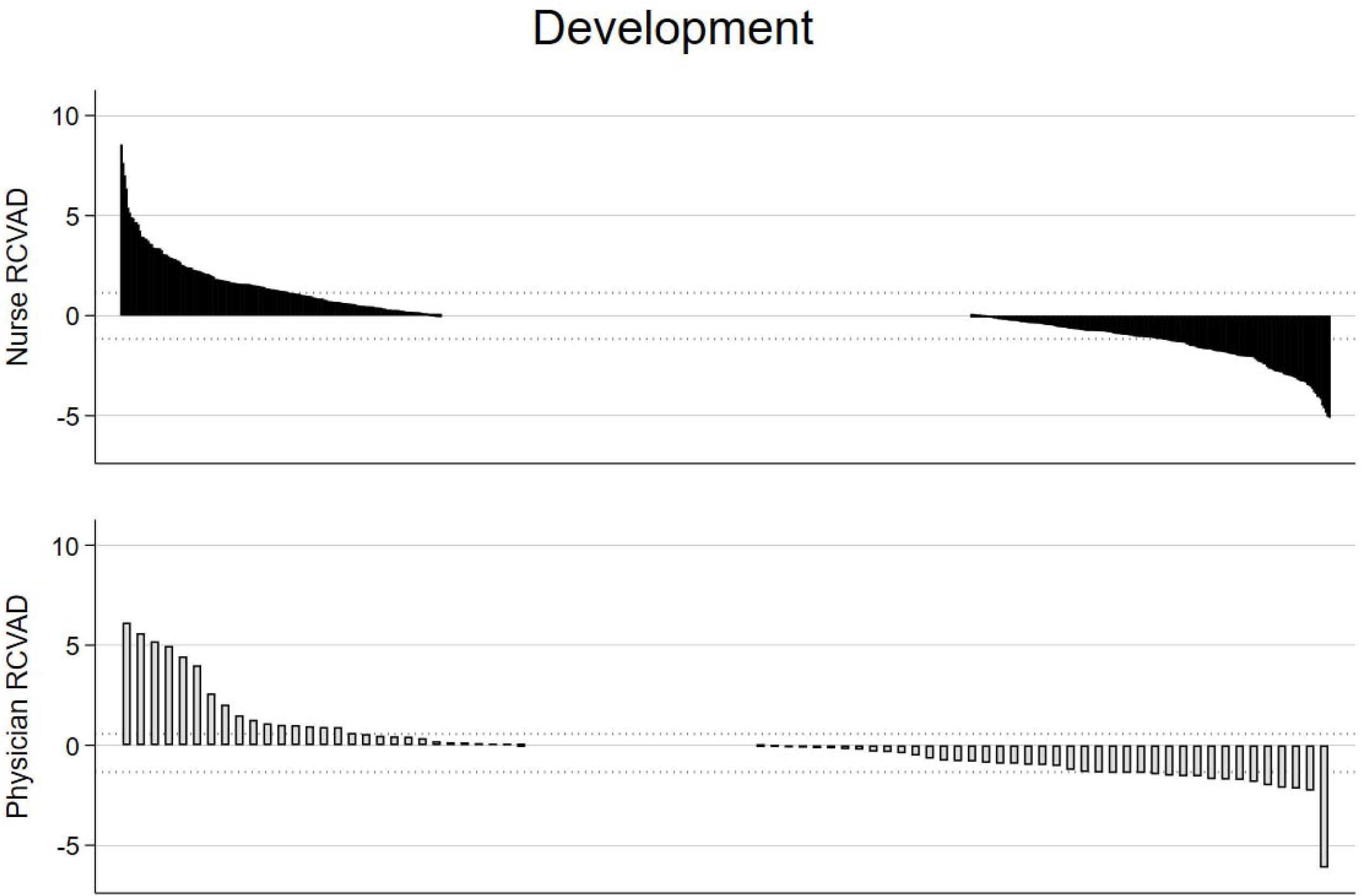
Clinician-specific race-contextual value-add difference. X-axis represents an individual clinician, ordered by decreasing race-contextual value-add difference (e.g., more disparate care delivered to Black patients). Dotted line represents bounds of the interquartile range. Top panels- nurses; bottom panels- physicians. 1A- Development partition. 1B- Testing partition. RCVAD- Race-contextual value-add difference.

**Figure 1B.**
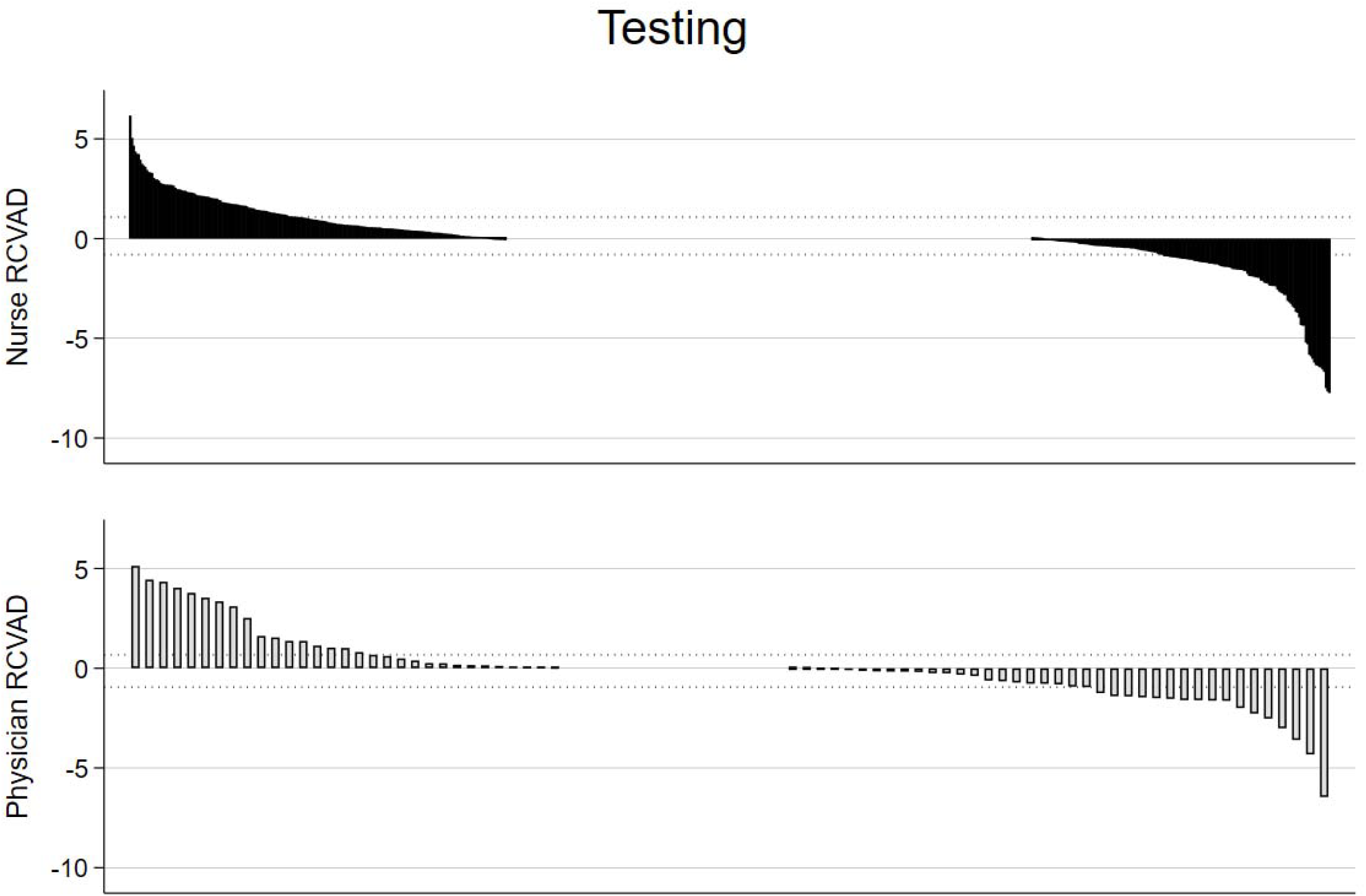
Clinician-specific race-contextual value-add difference. Dotted line represents bounds of the interquartile range. X-axis represents an individual clinician, ordered by decreasing race-contextual value-add difference (e.g., more disparate care delivered to Black patients). Top panels- nurses; bottom panels- physicians. 1A- Development partition. 1B-Testing partition. RCVAD- Race-contextual value-add difference.

**Table 2.** Summary statistics for the race-contextual value-add difference based on clinician type and partition.

| Clinician group | Partition | Mean | Standard deviation | Median | Interquartile range | Minimum | Maximum |
| --- | --- | --- | --- | --- | --- | --- | --- |
| Nurse | Development | 0 | 2 | -0.10 | (-1.17, 1.14) | -5.11 | 8.56 |
| Physician | Development | 0 | 2 | -0.18 | (-1.34, 0.56) | -6.12 | 6.14 |
| Nurse | Testing | 0 | 2 | 0.12 | (-0.80, 1.08) | -7.75 | 6.17 |
| Physician | Testing | 0 | 2 | -0.07 | (-0.95, 0.68) | -6.46 | 5.13 |

### Identifying assumption evaluation

*RCVAD* was positively correlated across both clinician groups (**Figure 2**; *r*=0.307, p<0.001 for nurses; and *r*=0.453, p<0.001 for physicians). *RCVAD* was not significantly correlated with the mean racial difference in LAPS at the time of first clinician assignment in either partition (**Figures S1-4**).

**Figure 2A.**
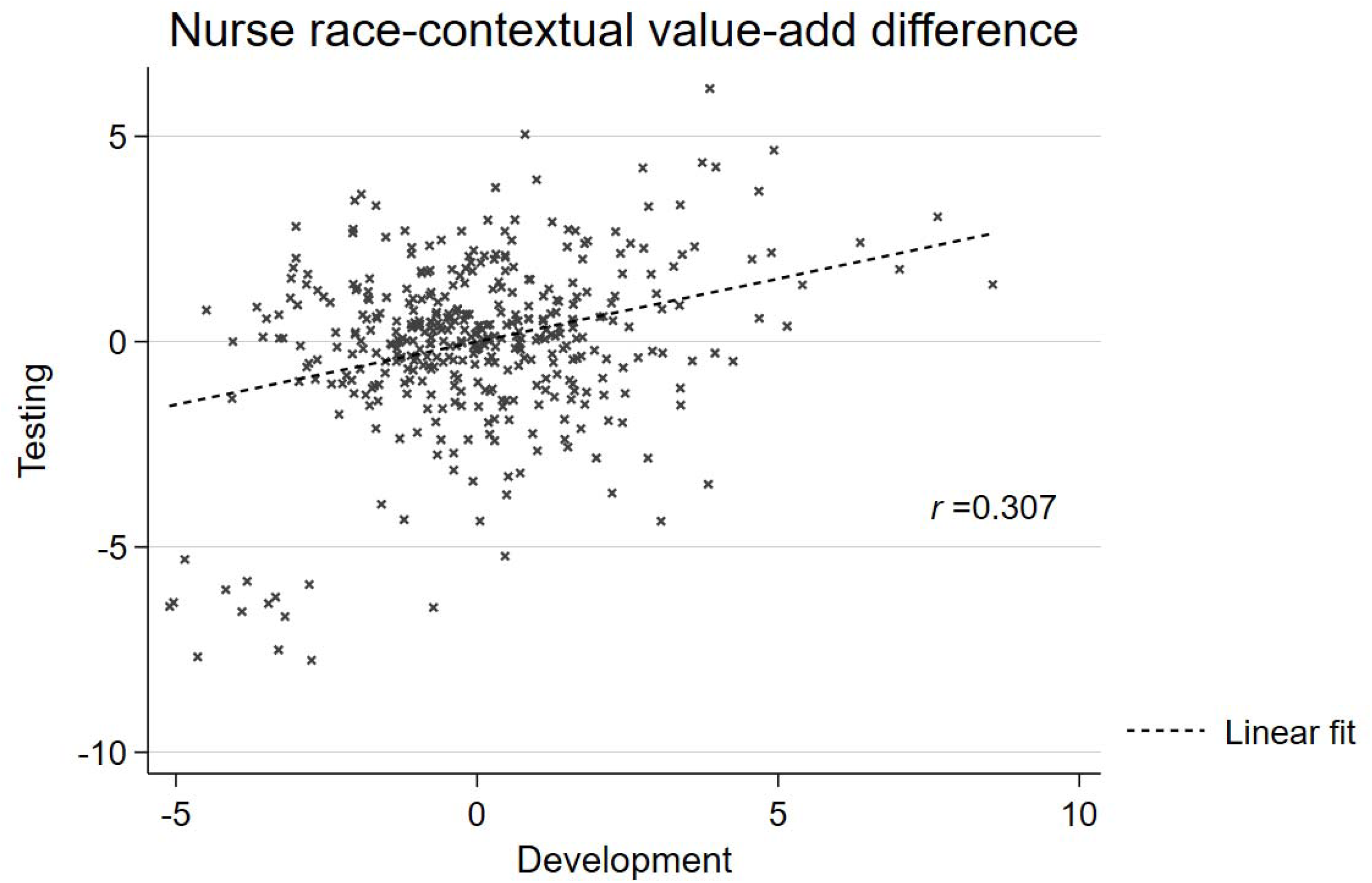
Scatter plot cross-validation of race-contextual value-add difference. 2A, nurses, 2B, physicians. *r*, Pearson correlation coefficient.

**Figure 2B.**
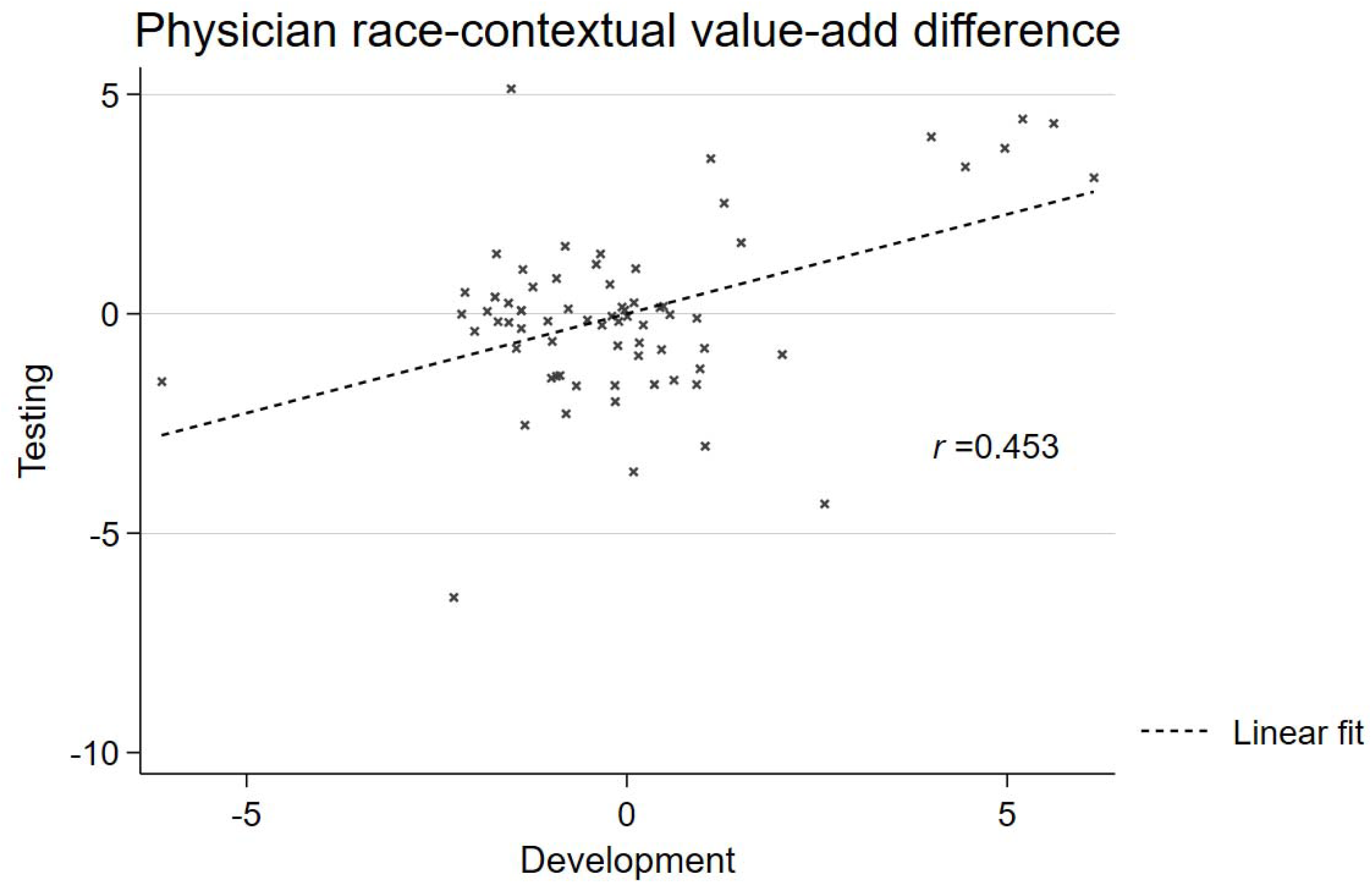
Scatter plot cross-validation of race-contextual value-add difference. 2A, nurses, 2B, physicians. *r*, Pearson correlation coefficient.

## Discussion

In this retrospective cohort, we demonstrated that Black patients experiencing mechanical ventilation not only were admitted with higher severity of illness than White patients, but also experienced more modest improvements in severity of illness during their ICU care relative to White patients. Additionally, we provide evidence that individual nurses and physicians are independently associated with both disparities-exacerbating and disparities-mitigating care with respect to severity of illness trajectory. Confidence in these findings stems from the fact that our analyses accounted for variation attributable to several patient-level, shift-level, and clinician-level fixed effects, including contemporaneous care provided by clinicians of either discipline. To accomplish this, we developed a novel measurement of clinician-level performance that differs with respect to patient race and that we define as the *RCVAD*. Because we demonstrate that the *RCVAD* fulfills key identifying assumptions, our approach is expected to reduce potential biases that may otherwise be introduced through value-added modeling approaches.

A second strength of this study is the racial diversity and sample size of clinical encounters. Third, because we define *RCVAD* as a measure of within-clinician differences in performance, unit- and hospital-attributable differences in resources and infrastructure are less likely to threaten *RCVAD* inferences than they might in conventional value-added modeling. Together, these findings provide evidence that the *RCVAD* reflects an independent measure of a clinician’s average difference in performance based on patient race. Additionally, because the *RCVAD* satisfies conventional VAM identifying assumptions, the *RCVAD* may be useful in future efforts to quantify independent clinician contributions to racially disparate clinical outcomes.

Moreover, this approach could also be extended to other fixed patient-level characteristics besides race, and could therefore evaluate how disparities based on rurality, gender, or other patient characteristics may stem, in part, from differential clinician performance.

In our approach, interpretation of 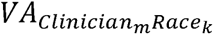 follows conventional value-add interpretation in which more positive values are associated with a greater improvement in severity of illness score (stated explicitly, a more negative Δ*LAPS*). This implies that a larger (or more positive) 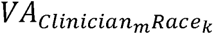 reflects higher performance than a smaller (or more negative) value. By inference, clinicians with *RCVAD* much greater than 0 would be expected to be associated with higher performance when caring for Black compared to White patients on average; the reverse would be true for clinicians with *RCVAD* much less than 0, who would be expected to be associated with higher performance when caring for White patients. As a result, one might infer that clinicians with *RCVAD* >0 have performance associated with disparities mitigating care, and those with *RCVAD* <0 have performance associated with disparities exacerbating care. In our cohort, large proportions of the clinician population were associated with potentially disparities-exacerbating care—up to 44-53% of nurses and 56-59% of physicians. An important caveat, however, is that the clinical impacts of the magnitude of *RCVAD* are unexplored, and should be characterized in future studies. Additionally, clinicians with a positive *RCVAD* still provide evidence of racially-discordant care practice and may have an opportunity to improve their average care delivery efficacy by improving care delivered to White patients.

Our approach has some limitations. First, the *RCVAD* may reflect multiple unstudied determinants of racially-discordant care delivery; this may include complex load balancing among clinicians and/or care delivery units, racially discordant availability of resources, or racially biased delivery of key resources. Our study was not designed to delineate which of these factors are strictly attributable to an individual clinician, and which factors influence a clinician’s care delivery; future studies might work to delineate the clinical delivery factors (including but beyond clinician characteristics) that function as mechanistic determinants for disparities mitigation and exacerbation. Second, our study took place in a single American academic center, limiting generalizability. Third, the demographic distribution of the study health system’s patient population and the use of effect modification in the *RCVAD* modeling approach limited our ability to quantify differences in clinician performance associated with patients other than Black or White race, limiting inferences to other groups. Fourth, our approach did not account for measures of socioeconomic disadvantage. Socioeconomic disadvantage may confound relationships between severity of illness at presentation and clinician performance since inpatient care delivery is associated with socioeconomic determinants such as insurance status.^16^ Lastly, cross-partition correlations were modest, and the possibility of residual confounding may affect the strength of associations that we measured.

## Conclusion

This study demonstrates that Black patients experiencing mechanical ventilation have less improvement in severity of illness compared to White patients during their ICU care and that the performance of individual nurses and physicians may influence differential trajectories in severity of illness. Because the performance of some clinicians suggests they may be exacerbating disparities while other clinicians may be mitigating disparities, future studies should seek to identify how these clinicians practice differently so as to enable more clinicians to perform as well when caring for patients of one race as another.

## Data Availability

Data are not available due to human subjects protections by the study institutional review board.

## Author Contributions

Study conception and design: CFC, MPK, RK, OY, and SDH. Data collection: SS and YL. Data analysis: CFC and YL. Writing of manuscript: CFC, MPK. Manuscript revisions for content and approval: all authors.

The authors state no conflicts of interest.

