## Supplement text for "Clinician contributions to disparities in severity of illness trajectories among mechanically ventilated patients"

Christopher F. Chesley, MD, MSCE

Olga Yakusheva, PhD

Yingying Lu, PhD

Rachel Kohn, MD, MSCE

Aerielle Belk, BS

Stefania Scott, MS

Scott D. Halpern, MD, PhD

Meeta Prasad Kerlin, MD, MSCE

**Online supplement**

|  | Unadjusted regression coefficient estimate (95% CI) | p-value | Adjusted regression coefficient estimate (95% CI) | p-value |
| --- | --- | --- | --- | --- |
| Black race (ref. White race) | 0.51 (-1.62, 1.64) | 0.639 | 2.26 (0.23, 4.29) | 0.029 |
| Patient age (years) | -- | | 0.25 (0.19, 0.31) | <0.001 |
| Female sex (ref. male sex) | -- | | -0.71 (-2.56, 1.13) | 0.447 |
| Elixhauser score | -- | | -0.02 (-0.07, 0.03) | 0.426 |
| Admission source (ref. ED) |  | |  |  |
| Direct | -- | | -15.88 (-18.75, -13.01) | <0.001 |
| Outside facility transfer | -- | | 6.41 (4.05, 8.77) | <0.001 |
| Surgical admission (ref. medical admission) | -- | | -3.64 (-5.74, -1.55) | 0.001 |
| Hospital duration prior to ICU admission (hours) | -- | | -0.02 (-0.02, -0.01) | <0.001 |
| Admission LAPS | -- | | -0.27 (-0.29, -0.25) | <0.001 |

**Table S1.** Adjusted association between the change in Laboratory Acute Physiology Score during ICU admission and patient race. Presented are regression coefficients from a generalized estimating equation model evaluating the relationship between the change in Laboratory Acute Physiology Score during ICU admission and patient race. Abbreviations: CI, confidence interval; ED, emergency department; ICU, intensive care unit, LAPS, Laboratory Acute Physiology Score 2; ref, reference.

**Supplemental Figures**

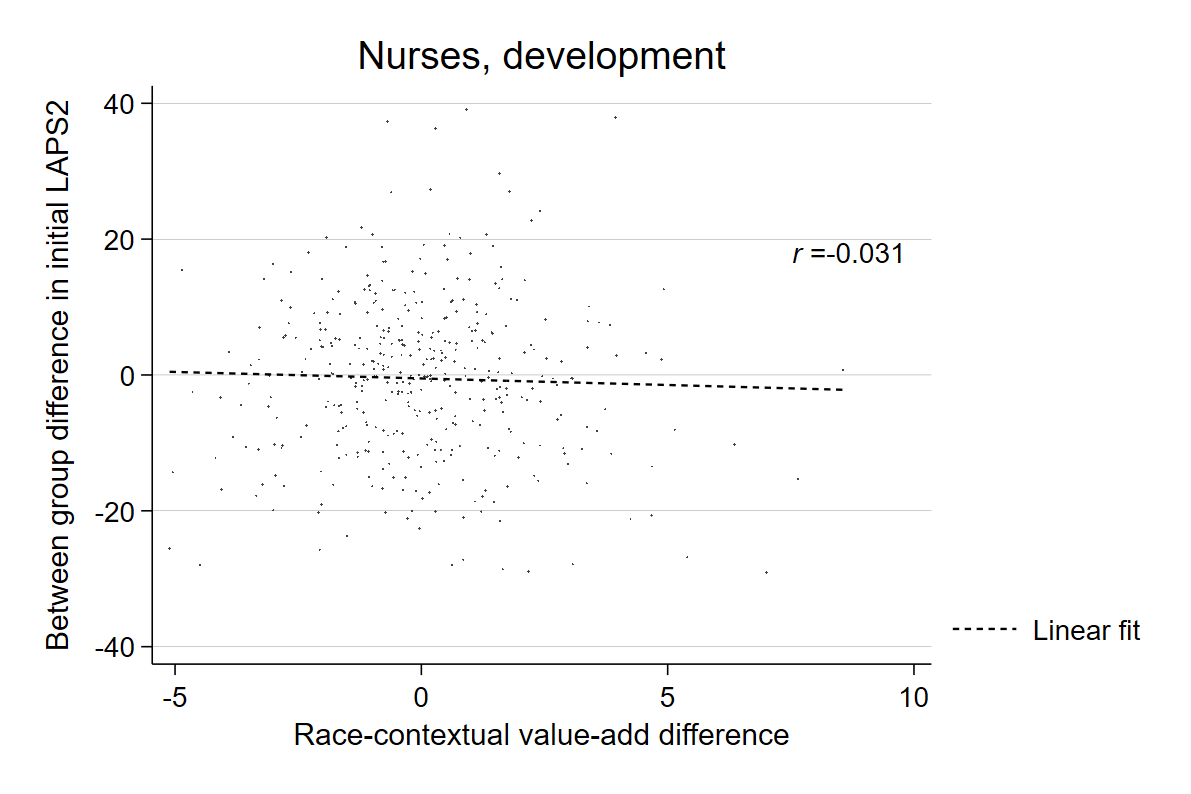

**Figure S1- Correlation between race-contextual value-add difference and mean race difference in severity of illness at time of first encounter-nurse assignment, development partition, p=0.537.**

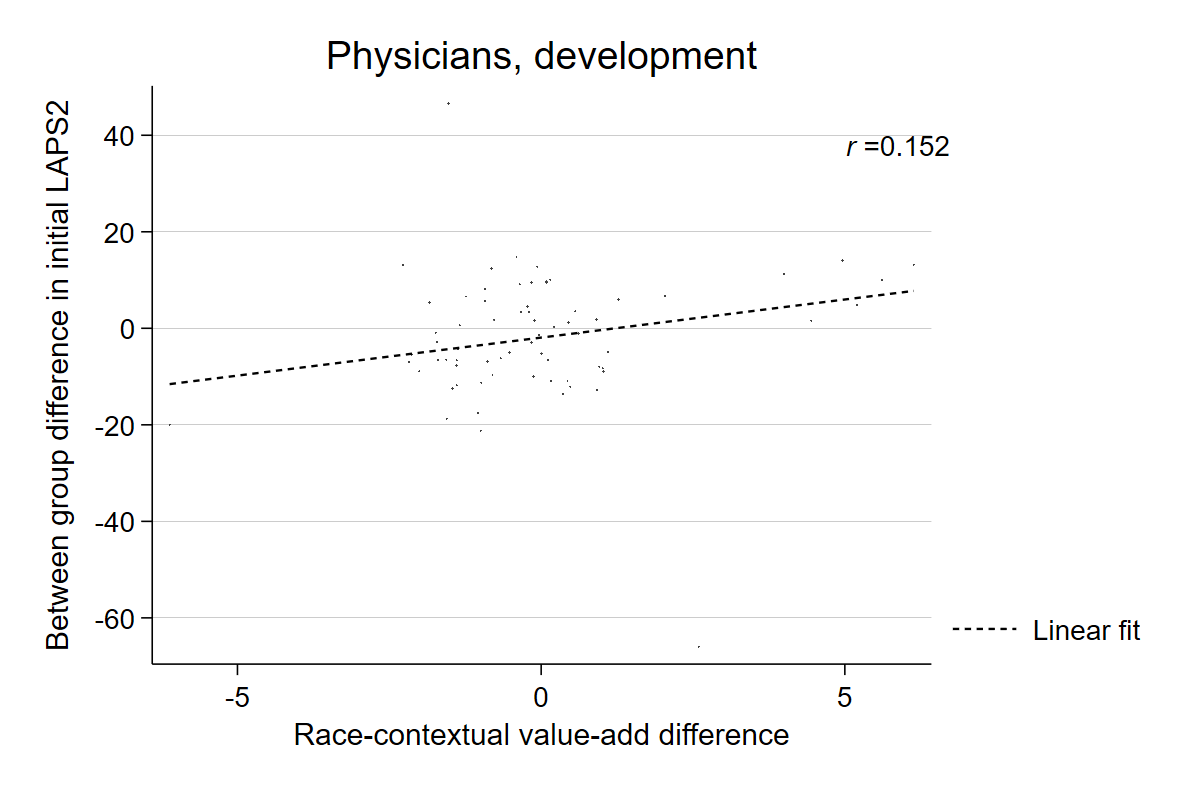

**Figure S2- Correlation between race-contextual value-add difference and mean race difference in severity of illness at time of first encounter-physician assignment, development partition, p=0.380.**

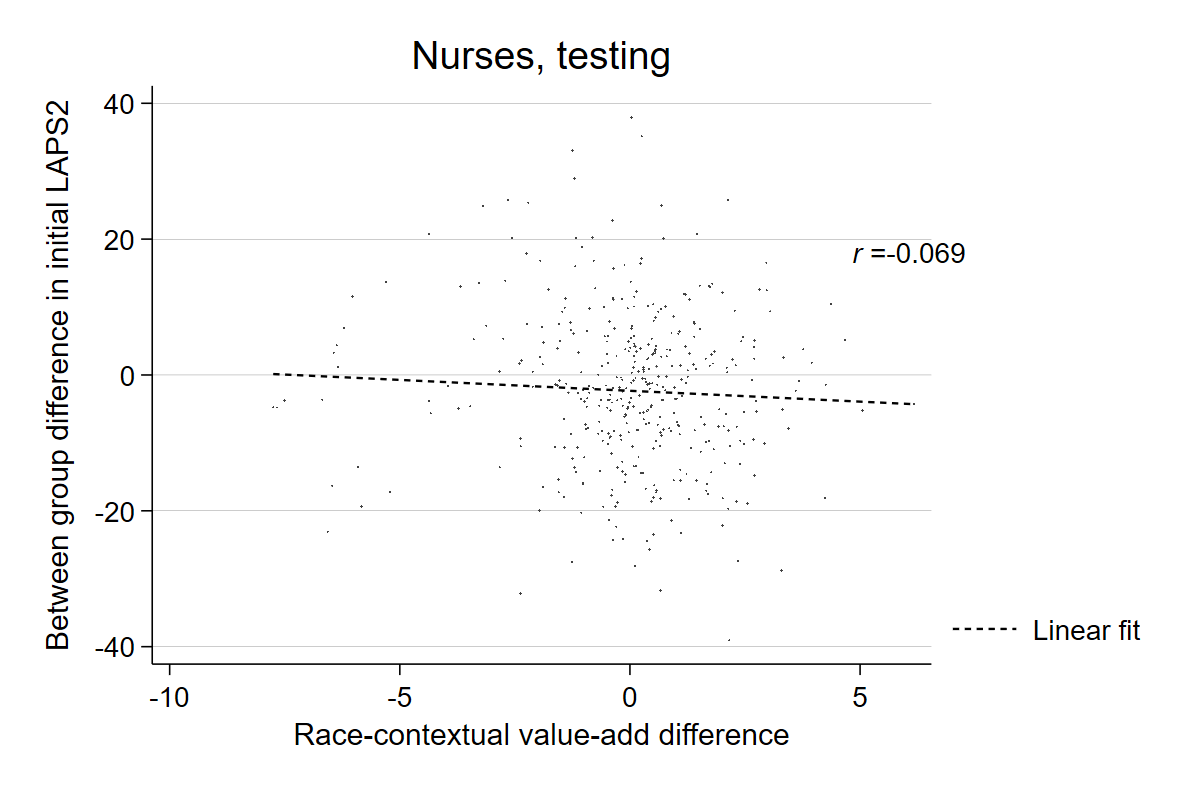
**Figure S3- Correlation between race-contextual value-add difference and mean race difference in severity of illness at time of first encounter-nurse assignment, testing partition, p=0.164.**

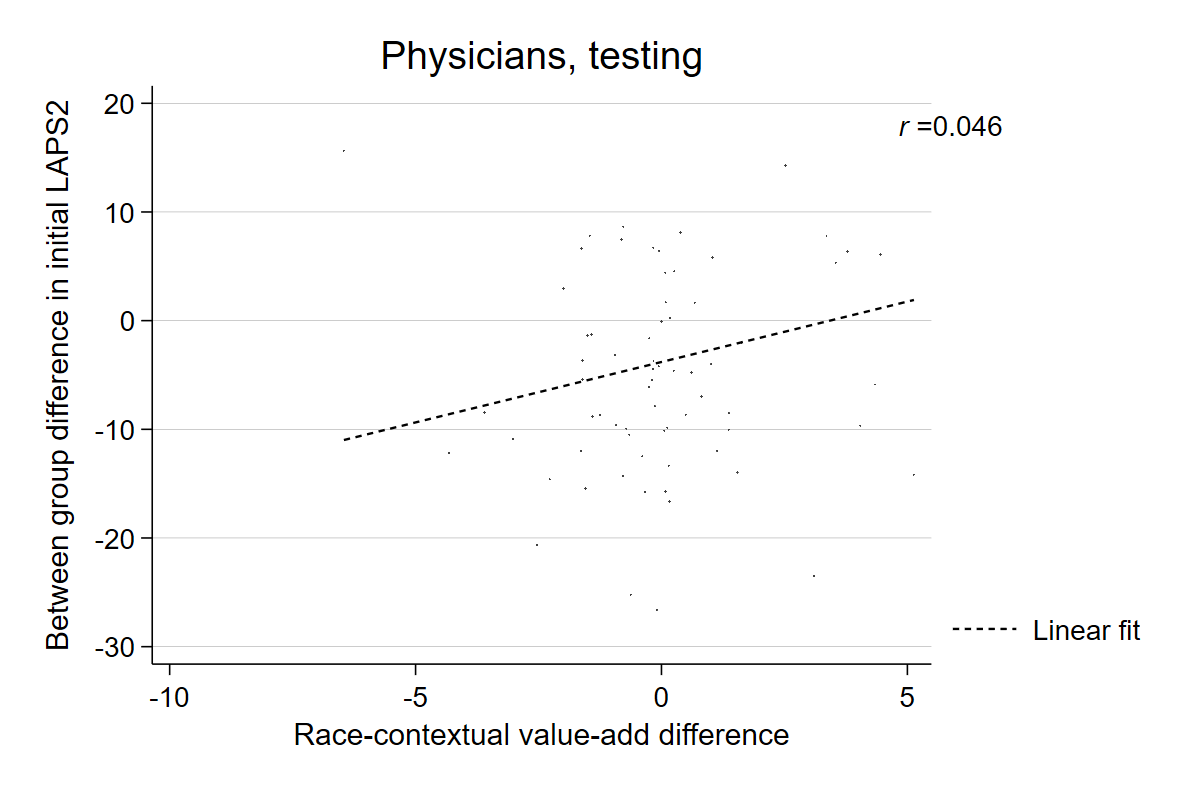
**Figure S4- Correlation between race-contextual value-add difference and mean race difference in severity of illness at time of first encounter-nurse assignment, testing partition, p=0.324.**
